# Efficacy and Outcomes of Pharmacological Treatments for Headaches after Traumatic Brain Injury: A Systematic Review

**DOI:** 10.1101/2025.08.11.25332976

**Authors:** Shirin Saleh, Elizabeth Sanford, Donald McGeary, Melissa Cortez, Matt Hayward, Erin D. Bouldin, Jacob Kean, Elisabeth A. Wilde, Mary Jo V. Pugh

**Affiliations:** Division of Epidemiology, Department of Internal Medicine, School of Medicine, University of Utah, Salt Lake City, UT, USA; Department of Psychiatry and Behavioral Sciences, University of Texas Health Science Center at San Antonio, San Antonio, TX, USA; Polytrauma Rehabilitation Center, South Texas Veterans Health Care System, San Antonio; Department of Neurology, School of Medicine, University of Utah, Salt Lake City, UT, USA; VA Salt Lake City Health Care System, Salt Lake City, UT, USA; Department of Population Health Sciences, School of Medicine, University of Utah, Salt Lake City, UT, USA; UT Health San Antonio Libraries, University of Texas Health Science Center at San Antonio, San Antonio, TX, USA

**Keywords:** Post-traumatic headache (PTH), Traumatic brain injury (TBI), Pharmacological treatment, Headache management, Efficacy, Systematic review, Quality of life, Randomized controlled trials (RCTs)

## Abstract

**Background/Objective:** Headache following traumatic brain injury (TBI) is a common, yet disabling, disorder with diverse mechanisms and treatment needs that remain poorly defined. Pharmacological regimens are the primary source of remedies for individuals with post-traumatic headaches (PTH). The main objective of this review is to describe the efficacy of pharmacological medications for the treatment of PTH with a specific focus on the effect of these medications on headache characteristics and patients’ quality of life (QoL).

**Methods:** This systematic review (CRD42024537710) followed PRISMA and SWiM guidelines. PubMed, CINAHL, Scopus, PsycINFO, and the Cochrane Library were searched in April 2024 for peer-reviewed articles published in English between 2009 and 2024. Eligible studies included randomized controlled trials, controlled cohort studies, and systematic reviews or meta-analyses evaluating pharmacological treatments for PTH in adults. Studies were excluded if they did not assess outcomes related to PTH pain, included pediatric populations, used animal models, investigated only non-pharmacological interventions, were case reports, narrative reviews, editorials, or conference abstracts, or did not involve human participants with TBI- or whiplash-related headache. Risk of bias was assessed using RoB-2 for RCTs and the Newcastle-Ottawa Scale (NOS) for controlled cohort studies.

**Result:** Sixteen studies were included in the final review. Most studies reported some improvements in headache frequency and intensity, with some also noting benefits in headache burden and QoL. Study designs included retrospective observational (n=7), non-randomized prospective (n=4), and randomized controlled trials (n=6). Of the RCTs, only three had a low risk of bias, and just one focused specifically on PTH.

**Conclusion:** Erenumab showed potential benefits for persistent headache symptoms and improved quality of life in civilian populations, while Prazosin demonstrated similar benefits in military populations, both with minimal side effects. Metoclopramide, co-administered with diphenhydramine to minimize side effects, demonstrated short-term efficacy as an abortive treatment for headache in emergency settings. However, due to the limited high-quality evidence, current pharmacological treatments for PTH should be used with caution. Future research should prioritize rigorous, controlled studies—particularly comparative effectiveness trials—and explore holistic, personalized approaches that incorporate treatment of psychiatric comorbidities and consider patient context.

---

Post-traumatic headache (PTH) is a common secondary headache disorder with onset following traumatic brain injury (TBI), whiplash, or similar physical injuries (1–7). According to the International Classification of Headache Disorders, 3rd version (8), PTH develops within seven days of the injury. If it resolves in 3 months, it is categorized as acute PTH, and it is considered persistent (PPTH) if the headache persists beyond three months (9). The prevalence of PTH ranges between 10% and 95% within the first year after TBI, influenced by factors such as population studied (e.g., athletes, military personnel, or general population), injury severity (10,11), and age and sex differences (12,13). In U.S. military populations, headache is the most common symptom after sustaining a TBI (14).

Acute PTH and PPTH are clinically complex conditions with a range of clinical presentations, which may complicate accurate diagnosis. Migraine (15–18) and/or tension-type headache (19–21) are among the most common clinical phenotypes of acute PTH and PPTH (22–24). Moreover, patients with PTH/PPTH often experience a range of physical symptoms, such as nausea, vomiting, photophobia, and phonophobia (25,26). Beyond the physical burden, PTH/PPTH is associated with impaired patient quality of life (QoL), daily functioning, and psychological well-being (27–29).

The American Academy of Neurology and American Headache Society (30) and U.S. Department of Veterans Affairs and Department of Defense (VA/DoD) Clinical Practice Guideline for the Management of Headache (2023 update) (31) provides recommendations for PTH management. The 2023 VA/DoD guideline additionally recommends newer pharmacotherapies and strategies that focus on value-based and patient-centered, tailored approaches. Pharmacological treatment is the primary approach for managing PTH in the acute phase. However, most pharmacotherapies prescribed for PTH/PPTH were developed for migraine or tension-type headache management (32–34). Medication options for these headaches are classified as preventative, intended to prevent the headache from occurring, compared to abortive, taken to terminate the headache in progress (35). Abortive medications like Non-Steroidal-Anti-Inflammatory Drugs (NSAIDs; e.g., aspirin, ibuprofen) are inconsistently effective (36). Preventative options that have been tested for PPTH, i.e., gabapentin, topiramate, and amitriptyline, aim to reduce the overall burden of headaches by decreasing their frequency and severity over time (33).

Many of the drugs prescribed for PTH/PPTH were developed for migraine and other primary headache conditions (37). The neuroinflammation model is widely used to provide a framework for developing pharmacotherapies to treat primary headache disorders (38,39). This model describes the cascade of inflammatory and cellular events that contribute to the development of chronic headaches through both peripheral and central mechanisms (40). **Figure 1** illustrates a generic pathway within this broader model, highlighting how some neuropeptides linked to headache pain (i.e., CGRP and PACAP-38) can lead to membrane depolarization and heightened pain sensitivity at the cellular level during migraine/headache. By contrast, PTH/PPTH results from a physical injury, not a primary headache disorder; thus, its pathways and presentation may not perfectly align with the neuroinflammation model (41). Nevertheless, many of the pharmacotherapies prescribed to manage PTH/PPTH—such as CGRP monoclonal antibodies (e.g., erenumab, fremanezumab), gepants (e.g., ubrogepant, rimegepant), NSAIDs, and investigational PACAP-38 inhibitors—are consistent with the neuroinflammation model.

**Figure 1.**
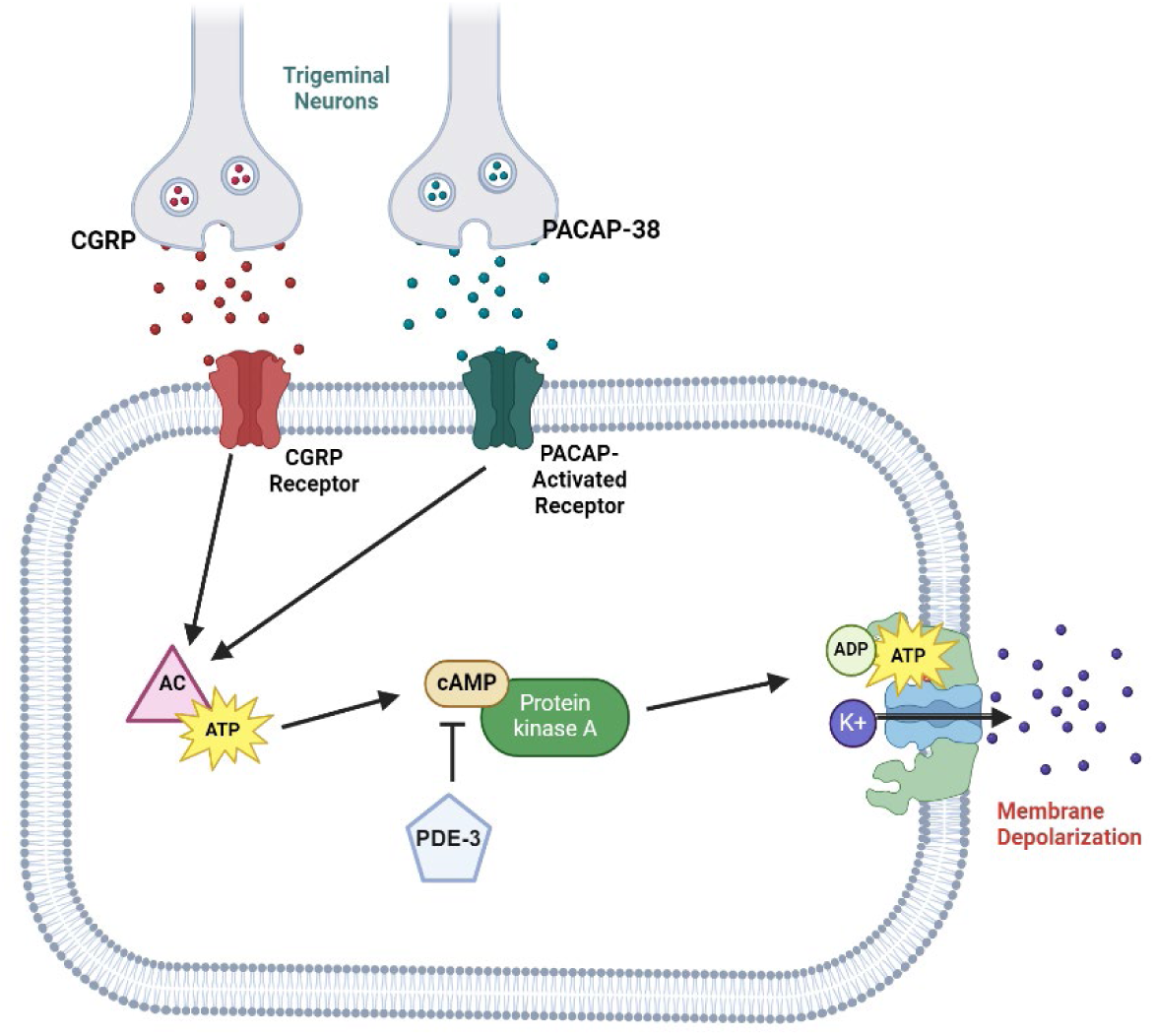
Potential Cellular-Level Pathways Associated with Headache Pain

The neuroinflammation model may, in large part, explain migraine or tension headache pain, but may not adequately explain PTH/PPTH pain. Interpreting headache subsequent to TBI through the lens of the neuroinflammation model assumes that exposure to neurotrauma alone may lead to chronic activation of the trigeminovascular system (14,23,33,42,43). Comorbid affective distress is common in people with TBI and may complicate treating PTH/PPTH (41). There may be evidence that comorbid affective distress states prolong or worsen pain (44,45). Service members with TBI, in particular, report a high incidence of clinically elevated depressive symptoms (46) and post-traumatic stress disorder (PTSD), which has been associated with headache-related disability in PTH/PPTH patients (47–49).

This review evaluates the available literature on pharmacological interventions used for the treatment of acute PTH and PPTH, exclusively in adults. Specifically, we evaluate the effectiveness of these treatments in reducing frequency, intensity, and headache-related disability, and in improving quality of life. Ultimately, this review aims to provide clinicians with a clearer understanding of the most effective pharmacological strategies for managing acute phase PTH and PPTH and contribute to refining current clinical guidelines to improve patient care.

## Methods

### Search Strategy

This systematic review is registered on PROSPERO with the registration number CRD42024537710. Searches were conducted in April 2024 in five bibliographic databases: PubMed, CINAHL, Scopus, PsycINFO, and Cochrane Library. The search strategy was developed in coordination with a librarian and is detailed in **Appendix 1**. The search string combined keywords and subject headings related to PTH treatments. This systematic review followed Preferred Reporting Items for Systematic Reviews and Meta-Analyses (PRISMA) (50) and the Synthesis Without Meta-analysis extension (SWiM) (51). The search was subjected to PRESS peer review using a modified version of the PRESS document (43) and was then translated into all included databases.

Two investigators (SS and ES) initially screened articles by title and abstract, followed by a comprehensive full-text review to determine eligibility for inclusion. Web-based Rayyan (52) was used to screen and select studies that met the inclusion criteria. Two additional investigators (DDM and MJP) reassessed all selected articles. By consensus among the four investigators, article selection was finalized. Key methodological details, sample characteristics, including headache persistence and TBI severity, pharmacologic agents, and outcome measures were reported.

### Eligibility Criteria

We included peer-reviewed randomized controlled trials, controlled cohort studies, observational studies, and systematic reviews or meta-analyses published in English between January 2009 and April 2024 that evaluated pharmacological treatments for acute-phase PTH or PPTH in adults (≥18 years). Eligible studies had to report at least one outcome related to headache pain, burden, or QoL in participants with TBI- or whiplash-related headache.

We excluded studies that: (1) did not assess outcomes related to PTH pain; (2) included pediatric populations; (3) used animal models; (4) investigated only non-pharmacological interventions; (5) were case reports, narrative reviews, editorials, or conference abstracts; or (6) did not involve human participants with TBI- or whiplash-related headache.

### Synthesis Methods and Effect Estimates

SWiM (Synthesis Without Meta-analysis) guidelines (51) and the Effect Direction Plot method (53) were used to systematically summarize and group each study into conceptual outcome domains (e.g., headache frequency, intensity, or impact). Within each domain, the direction of effect was assessed by counting the number of outcomes showing improvement, deterioration, or no change. Domains showing at least 70% of outcomes with the same effect direction were reported; if the effect was rated less than 70%, the effect was considered conflicting or unclear.

### Quality and Risk of Bias Assessment

Version 2 of the Cochrane Risk-of-Bias Tool for Randomized Trials (RoB-2) (54) and the Newcastle-Ottawa Scale (NOS) for controlled cohort studies (55) were implemented to assess bias in controlled studies. (Observational and retrospective studies were assumed to have a high risk of bias.) Randomized controlled trials (RCTs) were evaluated for quality based on randomization process, deviations from the intended interventions, missing outcome data, and outcome measurement (56). Controlled cohort studies were assessed based on the selection of participants, comparability of groups, and outcome measures. The direction and strength of treatment effects across all studies evaluating pharmacologic interventions for PTH/PPTH were summarized and categorized by outcome domains (headache frequency, intensity, disability, and QoL) and study design. Studies that reported positive effects were denoted using ▴ or ▴, to indicate the size of the effect. Studies indicating no change or mixed/conflicting results were denoted using ◀▶ (see **Table 1**).

**Table 1.** Effect Direction across Studies (Grouped by Risk of Bias)

| *Study* | *Design* | *Frequency* | *Intensity* | *Disability* | *QoL* | *RoB-2* |
| --- | --- | --- | --- | --- | --- | --- |
| *Ashina et al., 2020* | Contr Cohort | ▲<sup>2</sup> | ▲ | ▲ |  | High |
| *DiTommaso et al., 2013* | Contr Cohort |  |  |  |  | High |
| *Friedman et al., 2018* | Contr Cohort | ▲ | ▲ | ▲ | ▲ | High |
| *Ruff et al., 2009* | Contr Cohort | ▲ | ▲ | ▲<sup>2</sup> |  | High |
| *Charles, 2019* | Retro Obs | ▲ |  | ▲ |  | High |
| *Buture et al., 2023* | Retro Obs |  | ▲ | ▲ | ▲ | High |
| *Erickson, 2011* | Retro Obs | ▲ |  | ▲ |  | High |
| *Yerry et al., 2015* | Retro Obs | ▲ | ▲ | ▲ | ▲ | High |
| *Kazerooni et al., 2015* | Retro Obs | ▲ | ▲<sup>2</sup> |  |  | High |
| *Carabenciov et al., 2019* | Retro Obs |  | ▲ |  |  | High |
| *Hurwitz et al., 2020* | RCT | ◀▶ | ◀▶ | ◀▶ |  | High |
| *Cushman et al., 2019* | Retro Obs |  | ▲ | ▲ |  | Some |
| *Mayer et al., 2023* | RCT | ▲<sup>2</sup> |  | ▲ |  | Some |
| *Zirovich et al., 2021* | RCT | ▲<sup>2</sup> | ▲ |  |  | Some |
| *Friedman et al., 2021* | RCT | ▲ | ▲ | ▲<sup>4</sup> | ▲ | Low |
| *Hoffer et al., 2013* | RCT |  | ▲ |  | ▲ | Low |

Contr cohort = 'controlled cohort', non-randomized prospective study; Retro Obs = retrospective observational study; RCT = randomized controlled trial; QoL = quality of life; RoB-2 = Cochrane Risk-of-Bias Tool for Randomized Trials. Effect direction: upward arrow ▲ = positive effect, downward arrow ▼ = negative effect, sideways arrow ◀▶ = no change, mixed effects/conflicting results. Sample size in studies: Large Arrow ▲ > 300, medium arrow ▲ 50-300, small arrow ▲ < 50. Superscript numbers: Indicate the number of outcomes reported within each outcome domain. If not shown, the number is 1 by default. Study quality was visually incorporated using a traffic light color code: green=low risk of bias, amber=some concerns, and red= high risk of bias.

## Results

The database search yielded 2,280 search results, and two additional articles were identified through a manual search of reference lists of relevant primary articles. After removing duplicates, 1,693 articles were screened by title and abstract, and 21 were retrieved for full-text assessment. Ultimately, 16 articles met the inclusion criteria and are reviewed in this report (see **Figure 2**).

**Figure 2:**
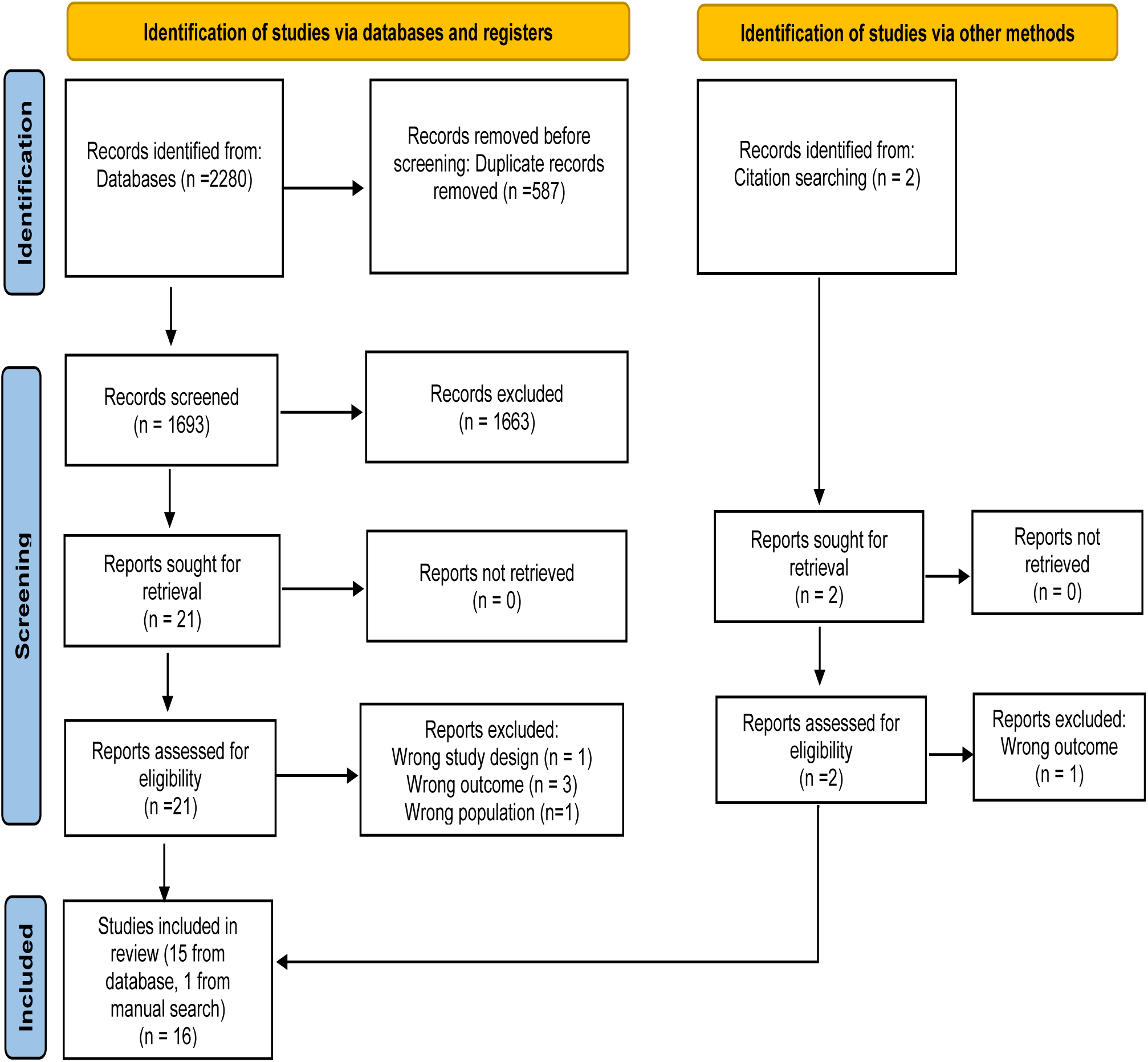
PRISMA Flow Chart Depicting the Selection Process of Studies for Systematic Review

Of the 16 studies, 10 specifically evaluated preventive medications (57–66), 4 studies focused on abortive treatments (67–70), and 2 combined both preventive and abortive treatments (71,72). Populations of interest among studies were highly heterogeneous: 9 articles included civilians with either acute or persistent PTH or adult athletes with a history of concussion (57–59,61,65,66,68,70,73), while the remaining seven used samples drawn from military service members and/or veteran populations with either acute phase PTH or PPTH (60,62–64,67,71,72). Fourteen out of 16 studies (87.5%) reported on samples with mTBI, and the other two studies did not specify severity. Five were RCTs with a placebo-control (60–62,67,73), four studies used a controlled cohort design, and the remaining seven studies were observational (57–59,63–66,70–72). Most showed a high risk of bias (11 out of 16, 70%), with only 2 studies rated as low risk. Controlled cohort and oobservational studies were reported in **Table 2**. RCTs were reported in **Table 3**.

**Table 2.** Overview of the Observational Studies on Pharmacological Headache Treatment.

| Source | Study Design | Study Cohort<br>Baseline<br>Characteristics | Intervention<br>(Dose/Type/Adverse<br>Events/) | Outcome<br>Measures | Desired Effects (Changes<br>in intensity,<br>frequency, duration) | Findings/<br>Key Points |
| --- | --- | --- | --- | --- | --- | --- |
| **Ashina et al., 2020** | Non-randomized, single-arm clinical trial<br>Screening phase: 0 to 2 weeks; Baseline phase: 4 weeks; Open-label treatment phase: 12 weeks<br>Scheduled visits at: Screening; Baseline (dose 1); Week 4 (dose 2); Week 8 (dose 3); Week 12 (final evaluation) | n= 100, Civilian<br>75% F, 25% M<br>Mean age: 35.1<br>PPTH<br>HA phenotype: Chronic migraine-like (53%), combined episodic migraine-like/TTH-like HA (34%), pure chronic TTH-like (13%) | Erenumab<br>140 mg monthly by two subcutaneous 1-mL injections<br>Preventative<br>AE: 78 patients; common reports: constipation (30), injection-site reactions (15), dizziness (9), worsened HA (8) | Reduction in headache frequency and intensity<br>Proportion of patients achieving $\geq 25\%$, $\geq 50\%$, or $\geq 75\%$ reduction in headache days<br>HIT-6 | $2.8 \pm 6.8$-day reduction in moderate to severe HA by weeks 9-12<br>28% achieved $\geq 50\%$ reduction in mean number of moderate to severe HA days | Erenumab reduced the frequency of moderate to severe HA days in patients with PTH<br>Well-tolerated: 89% of patients received all three doses<br>Low discontinuation rate due to adverse events |
| **Charles et al., 2019** | Retrospective case series<br>Pre -post comparison<br>2 months FU indicated no relapse | n=7, Civilian<br>F=5, M=2<br>Mean age: NS<br>Acute PTH<br>HA phenotype: Migraine | Erenumab<br>140 mg x1<br>Preventative<br>AE: low incidence of injection site reactions and constipation | Baseline MHDs<br>HIT-6 | 95% reduction in the mean number of HA days (SD 1.22, $p < .001$) | Effective in treating PTH with migraine phenotype |
| **Buture et al., 2023** | Retrospective, open-label, observational audit in a real-world setting<br>Measures collected every 3-12 months until study ended in 2019 | n=82<br>F=54, M=28<br>F mean age: $44 \pm 13$<br>M mean age: $45 \pm 14$<br>PPTH<br>HA phenotype: Treatment-refractory headache conditions | Erenumab<br>70 mg for 3-4 months; increase to 140 mg if inadequate response/no side effects; 21% started at 140 mg (BMI $>30$)<br>Preventative<br>AE=NS | HIT-6<br>MIDAS<br>MSQ | 29 (35%) improved headache intensity and migraine symptoms<br>Median reduction in HIT-6 (4.25-5 points) over the first year<br>Improvement in MIDAS scores compared to baseline<br>Reduction of 14 points in the MSQ scores (improved QoL) | 35% reported subjective improvement in HA severity and migraine symptoms<br>65% stopped treatment within 6-25 months mainly due to lack of efficacy, or side effects<br>Erenumab offers a meaningful benefit in a subset of highly refractory patients |
| **Erickson, 2011** | Retrospective cohort study<br>FU outcomes via standardized phone | n=100 (77 Blast, 23 non-blast), Active-duty US Army<br>F=1, M=99<br>Mean age: 28.7 | Amitriptyline 25-50 mg, Nortriptyline 25-50 mg (antidepressants);<br>Topiramate 100 mg, | Headache frequency<br>MIDAS | HA frequency reduced from 17.1 to 14.5 days /month ($P = .009$)<br>Non-blast PTH patients showed a greater reduction in | Topiramate effective for PPTH in military<br>Triptans are often effective for aborting acute headaches in PPTH |
|  | questionnaire at 3 months post-baseline | • PPTH<br>• HA phenotype: Mostly migraine | Propranolol LA 80 mg, Valproate ER 500 mg (prophylactic); Triptans<br>• Abortive<br>• AE=NS | • Response to abortive medications | headache frequency (41%) compared to blast PTH patients (9%)<br>• Topiramate users: 23% reduction in HA frequency (P = .02)<br>• Triptan users: 70% relief within 2 hours (P = .01) | • Low-dose TCAs show little efficacy<br>• Blast-related PTH may be less responsive to prophylactics |
| **Ruff et al., 2009** | • Observational study with pre- and post-assessment<br>• FU:9-week and 6-month | • n=74, Veterans from Cleveland OH<br>• F=5 M=69<br>• Mean age: 29.4 (SD=2.9)<br>• PPTH<br>• HA phenotype: 40.5% TTH, 18.9% migraine-like HA, and 40.5% mixed-type HA | • Prazosin + Sleep hygiene counseling<br>• Dose started at 1 mg QHS (at bedtime) and increased weekly to 2 mg, 4 mg, 5 mg, and 7 mg. Final treatment dose was 7 mg at bedtime<br>• Preventative<br>• AE= Abnormalities in liver function, changes in bowel habits, skin rash or pruritis, daytime somnolence, light-headedness, postural hypotension | • Headache pain intensity (NRS)<br>• Headache frequency per month<br>• MOCA<br>• Sleep quality | • Peak HA pain decreased from 7.28 (SD = 0.27) to 4.08 (SD = 0.19)<br>• HA per month reduced from 12.40 (SD = 0.94) to 4.77 (SD = 0.34)<br>• Cognitive performance improved from 24.5 (SD = 0.49) to 28.6 (SD = 0.59) after 9 weeks | • Sleep gains from prazosin maintained at 6 months<br>• Improving sleep is an effective first step for treating PPTH in OIF/OEF veterans |
| **Cushman et al., 2019** | • Retrospective study with 3 groups(no meds, gabapentin, TCAs)<br>• 1-year FU<br>• Mixed effects analysis for group comparisons | • n=277, Civilian/athletes<br>• No-Med group (n=123), M: 40.3%, F: 48.5%, Mean age: 20.9 (SD=12.4)<br>• Gabapentin group (n=60), M: 25.9%, F: 17.4%, Mean age: 24.8 (15.6)<br>• TCA group: (n=94), M: 33.8%, F: 34.1%, Mean age: 24.6 (14.6)<br>• PPTH<br>• HA phenotype: NS | • TCAs and gabapentin<br>• Median doses: Gabapentin 900 mg, TCAs 20 mg<br>• Preventative<br>• AE= cognitive side effects, including drowsiness, mental slowness, and nausea | • PCSS | • HA dropped from 4.4 to 3.2 after TCAs, not significant<br>• Symptom score decreased significantly with TCAs (64 to 33, p = .005)<br>• No significant symptom decreases with gabapentin | • HA and SY burden showed modest improvement over time<br>• More sustained improvement with TCAs compared to gabapentin |
| **Yerry et al., 2015** | Real-time retrospective consecutive case series between August 2008 to August 2012<br>Charts reviewed using pharmacy records searched by ICD-9 code for chronic migraine and PTH | n=64, Active-duty service members<br>F=1 M=63<br>Mean age: 31.3 (SD=7.5)<br>PPTH<br>HA phenotype: mixed (chronic TTH/migraine/ CD) | BoNT/A<br>FDA FSFD (155U) was the primary protocol; some received FSFD+CD or FSFD+FTP (~200U); a few had FTP only or focal injections (e.g., 50U for nummular pain)<br>Preventative<br>AE= neck pains and HA | GEC<br>Occupational outcome<br>HA characteristics | GEC improved in 41 (64.1%), unchanged in 18 (28.5%), worsened in 2 (3.2%)<br>Of 48 soldiers with continuous HA, 26 (54.1%) stopped treatment, 11 (22.9%) experienced some or much improvement, 22 (46%) left the military | 2/3 of patients reported improvement with BoNT/A<br>Patients treated further from injury more likely to remain on active duty<br>BoNT/A offers a well-tolerated preventive option for PPTH<br>1/3 continued treatment; main reason for discontinuation was lack of improvement |
| **DiTommaso et.al., 2013** | Observational cohort study<br>Patients in level 1 trauma center in Seattle<br>FU at 3, 6, 12 months post injury | n= 212, Civilian (analysis focused on the 167 with HA symptoms)<br>F (25%) M (75%)<br>Mean age: 42.1<br>Acute and PPTH<br>HA phenotype: Migraine, TTH, cervicogenic and unclassified | Self-reported medication use patterns rather than administering specific interventions<br>Most commonly NSAIDs and acetaminophen, but also triptans, opiates, antiseizure, anxiolytics and antidepressants<br>Doses varied by medication type<br>AE= NS | Self-reported effectiveness of headache treatment by participants following mTBI | Most participants reported that their medications provided partial or no relief.<br>Only 26% of those with migraine/probable migraine and 70% with TTH reported complete relief. | Overall, headache treatment was generally inadequate, especially for migraine phenotypes. |
| **Kazerooni et al., 2015** | Retrospective case series with electronic chart reviews<br>Typical FU at 3 months after initial injection | n=21, US veterans<br>F= 52%, M=48%<br>Mean age: 40 (SD=9.5)<br>PPTH<br>HA phenotype: Migraine | Incobotulinumtoxin A<br>150 units of incobotulinumtoxin A per injection cycle<br>Preventative<br>AE= NS | Headache days/month<br>Headache intensity<br>Duration of action | Significant reduction in HA days per month: 19.1 vs. 9.1 days ($P < 0.001$)<br>Significant reduction in HA intensity: 8.3 vs. 4.1 ($P < 0.001$)<br>Those with a history of TBI, HA days/month decreased from 19.5 to 6.7 | Duration of action averaged 81.9 days (median 70 days)<br>IncobotulinumtoxinA was effective in a small patient population |
| **Carabenciov et al., 2019** | Retrospective review of consecutive patients with PCS at Mayo Clinic<br>No comparison group or placebo<br>33% dropout rate before completing a full course | n=33 (11 discontinued for AE; 22 completed a full trial), Civilian<br>F=13 M=20<br>Mean age: 35<br>PPTH<br>HA phenotype: Mixed presentation | Amantadine<br>Complete trial was defined as 100 mg twice per day for 2 months<br>Preventative<br>AE= 11 discontinued for dizziness, nausea, and/or worsened HA | Symptom improvement based on physician documentation | 80% of patients (n=22) had improvement in their HAs. | Amantadine was prescribed for PCS, not specifically PTH<br>80% of patients who completed the full trial (n=22) experienced HA improvement |
| **Friedman et al., 2018** | Prospective, open-label study for patients with acute PTH in the ED<br>FU at 1 hour, 2 hours, 48 hours, and 7 days for some outcomes | n=21, Civilian<br>F=16 M=5<br>Mean age: 45<br>Acute PTH<br>HA phenotype: NS | IV metoclopramide + diphenhydramine<br>Metoclopramide 20 mg + diphenhydramine 25 mg across 15 minutes<br>Abortive<br>AE= sleepiness | Ordinal pain scale<br>Functional status<br>Patient satisfaction<br>PCSS<br>SCAT<br>Headache frequency<br>Return visits to healthcare providers | 95% reported HA improvement to no worse than mild<br>Mean PCSS Score improved from 47.5 (SD 29.4) before treatment to 10.9 (SD 14.8) at ED discharge<br>Score remained at 11.4 (SD 21.4) one week after treatment | IV metoclopramide 20 mg + diphenhydramine 25 mg is effective and well-tolerated for acute PTH in ED<br>2/3 report no relapse after discharge<br>75% report continued relief one week later |

F females, M males, HA headache, PTH post-traumatic headache, PPTH persistent post-traumatic headache, mTBI mild traumatic brain injury, AE adverse events, TTH tension-type headache, FU follow-up, HIT-6 headache impact test-6, MIDAS migraine disability assessment, MSQ migraine specific quality of life, MHDs monthly headache days, SD standard deviation, mg milligram, NS not specified, OIF operation Iraqi freedom, OEF operation enduring freedom, BMI body composition index, TCAs tricyclic antidepressants, MOCA Montreal cognitive assessment, NRS numeric rating scale, SY symptom, ICD-9 international classification of diseases 9th edition, BoNT/A onabotulinum toxin A, FDA food and drug administration, FSFD fixed site, fixed dose, CD cervical dystonia, FTP follow the pain, U units, GEC global evaluation of change, NSAIDs nonsteroidal anti-inflammatory drugs, PCS post-concussion syndrome, ED emergency department, IV intravenous, PCSS post-concussion symptom scale, SCAT sport concussion assessment tool

**Table 3.** Overview of the Clinical Trials on Pharmacological Headache Treatment.

| Source | Study Design | Study Cohort<br>Baseline<br>Characteristics | Intervention<br>(Dose/Type/Adverse Events/) | Outcome Measures | Desired Effects (Changes<br>in intensity,<br>frequency, duration) | Findings/<br>Key Points |
| --- | --- | --- | --- | --- | --- | --- |
| **Mayer et al., 2023** | Parallel-group RCT, 22 weeks total<br>4-week baseline, 5–7-week dose titration, 12-week maintenance<br>2:1 prazosin to placebo ratio<br>Completion rates: prazosin 94%, placebo 88% | n=48, Veterans and Active duty US service members<br>F=44, M=4<br>Mean age: 43<br>PPTH<br>HA phenotype: Migraine, TTH, and mixed | Prazosin<br>Titrated gradually upward; maximum dose of 5 mg morning and 20 mg evening, or as tolerated,<br>Preventative<br>AE= Dizziness/vertigo, Syncope/fall, Palpitations, Nasal congestion, Nausea/vomiting, Morning drowsiness/lethargy, Peripheral edema, Urinary incontinence, Hypotension | Change in the 4-week frequency of qualifying headache days<br>Reduction in qualifying headache days<br>Change in HIT-6 scores | Baseline HA: prazosin 18.3 (±1.6), placebo 18.6 (±2.3) per 4 weeks<br>At week 12, prazosin group had $11.9 \pm 1.0$ fewer HA days vs. $6.7 \pm 1.5$ for placebo (5.2 days difference, 95% CI [−8.8, −1.6], $p = 0.005$)<br>$\geq 50\%$ reduction in HA frequency ($p = .004$)<br>Prazosin group had lower HIT-6 scores ($p = .004$) | Study drug was generally well tolerated |
| **Zirovich et al., 2021** | Prospective, double-blind, randomized, placebo-controlled, cross-over trial<br>Monthly FU for 11 weeks | n=40, Veterans from LA, CA<br>F=2 M=38<br>Mean age: ~34<br>PPTH<br>HA phenotype: Mixed migraine and TTH | BoNT/A<br>0.1 mL of either BoNT/A or normal saline was injected into 31 facial and cervical muscle sites, including the frontalis, corrugator, procerus, temporalis, occipitalis, cervical paraspinals, and trapezius, as per BoNT/A clinical trials<br>Preventative<br>AE= pain, forehead paresthesia, itching at the injection site, sinusitis, dizziness, blurred vision, eyelid ptosis, eye irritation, neck weakness, sensitivity to light, and twitching eye | Headache frequency<br>Headache days per week<br>Headache pain severity | HA per week decreased by 2.24 (43.3%) with BoNT/A ($P < .001$)<br>HA days per week decreased by 2.24 (44.4%) at 16 weeks ($P < .001$)<br>HA pain severity reduced by 0.06 with BoNT/A ($P = .02$) | Significant reduction in number of HA days/week in the BoNT/A group compared to placebo over 16 weeks<br>HA pain significantly reduced<br>AE were generally mild and transient, typical of injection site responses |
| **Hurwitz et al., 2020** | • Single-center, 3-month, 2-arm (immediate vs. delayed) prospective phase II trial<br>• Assessing efficacy and feasibility<br>• 90-day duration | • n=50 (24 immediate, 26 delayed), Civilian<br>• F=22 M=28<br>• Mean age: 35.7 (SD=12.2)<br>• PPTH<br>• HA phenotype: NS | • Amitriptyline<br>• Gradually increased from 10 to 50 mg daily<br>• Preventative<br>• AE= Increased QTc, dry mouth, decreased libido, daytime fatigue, increased HA | • Headache frequency<br>• Headache intensity<br>• Medication compliance<br>• Diary compliance<br>• Cognitive testing | • Low compliance with daily HA diary: 58% (29/50) completed daily entries<br>• Only 10 participants had 100% completion<br>• No differences in HA frequency or severity between immediate and delayed medication start | • Could not determine the benefit of amitriptyline as a HA preventive due to challenges with recruitment, retention, and compliance |
| **Friedman et al., 2021** | • Randomized, double-blind, placebo-controlled, parallel-group study<br>• FU at 48 hours and 7 days later | • n=160 (79 placebo, 81 exp), Civilian<br>• F=107 M=53,<br>• Mean age: Placebo=43, exp=6<br>• HA phenotype: Acute PTH | • IV metoclopramide + diphenhydramine<br>• Metoclopramide 20 mg + diphenhydramine 25 mg across 15 minutes<br>• Abortive<br>• AE= drowsiness, akathisia, dizziness, diarrhea, and typical PCS symptoms | • Improvement in headache intensity<br>• Sustained headache relief over 48 hours<br>• PCS measured by the PCSS<br>• Use of rescue medication<br>• Patient satisfaction<br>• Headache frequency | • Placebo group reported a mean improvement of 3.8 (SD 2.6)<br>• M + D group improved by 5.2 (SD 2.3)<br>• Difference favoring metoclopramide was 1.4 (95% CI 0.7–2.2, p < 0.01) | • M + D was more effective than placebo for relief of PTH in the ED |
| **Hoffer et al., 2013** | • Randomized, double-blind, placebo-controlled at a forward deployed field hospital in Iraq<br>• Evaluations at 3- and 7-days post-baseline | • n=81 (Placebo = 40, NAC=41), Service members<br>• F=1 M=80,<br>• Mean age: 22 (range 18-43 years)<br>• Acute PTH<br>• HA phenotype: NS | • NAC<br>• 4 grams NAC/day (8 x 500mg pills)<br>• Abortive<br>• AE= NS | • Complete symptom resolution by Day 7<br>• Improvement in neuropsychological performance (TMT, COWA, AN) | • Subjects receiving NAC within 24 hours of blast had an 86% chance of symptom resolution<br>• No reported side effects with NAC<br>• Compared to 42% symptom resolution for placebo | • NAC is an effective short-term countermeasure for cognitive and PCS symptoms from blast damage<br>• Early treatment within 24 hours of blast exposure shows greater efficacy |

RCT Randomized Controlled Trial, BoNT/A botulinum toxin type, PTN posttraumatic neuralgia, PGIC patient global impression of change, NPSI neuropathic pain symptom inventory, TMT trail making test A & B, COWA controlled oral word association, AN animal naming, QTc corrected QT interval

### Preventative medications

#### Monoclonal antibody treatments

Three studies investigated the efficacy of erenumab, a CGRP receptor monoclonal antibody therapeutic, as a preventative medication for PTH/PPTH. CGRP is associated with increased pain sensitivity (**Figure 1**). CGRP has been shown to exacerbate headaches with migraine-like features in patients with PPTH (74).

Charles et al. (2019) conducted a retrospective case series (n=7) to evaluate the effect of erenumab in the acute phase of PTH after sustaining mTBI. Patients who met clinical criteria and had failed at treatment or were intolerant of conventional migraine preventatives (58). The medication was generally well-tolerated and showed effectiveness within days to four weeks, with most requiring only one dose. The study provided preliminary evidence suggesting that erenumab may be effective for treating PPTH. However, the findings should be interpreted with caution: there were only a few cases, there were no controls or comparators, and assessments were retrospective.

Buture et al. (2023) assessed erenumab among patients (n=82) with treatment-refractory headache conditions, including those diagnosed with PPTH following minor head trauma or other rare secondary headache disorders (59). Of the 15 patients diagnosed with PPTH, 9 showed a meaningful response to erenumab treatment, suggesting there may be utility for exploring it as a long-term preventative. However, no conclusions should be drawn without an RCT.

Ashina et al. (2020) conducted an open-label study evaluating the effectiveness of erenumab for PPTH following mTBI (57). Seventy-four percent (n=100) of patients had a history of preventive medication use, and 86% experienced failure with at least one previously attempted preventive treatment. Patients reported a significant reduction in the frequency of moderate to severe headache days per month by week 12, with minimum adverse events. Short-term effects only were evaluated, limiting conclusions regarding long-term safety and efficacy. The findings suggest erenumab is promising as a short-term preventive treatment for PPTH. RCTs are urgently needed to draw any further conclusions.

#### Sleep Aids

Two studies investigated the efficacy of prazosin, an antihypertensive agent, for PPTH. Prazosin is commonly used off-label to manage sleep problems related to PTSD symptoms, particularly nightmares and sleep disturbances (75,76). By targeting alpha-1 receptors in the brain, prazosin reduces the hyperarousal that contributes to nighttime PTSD symptoms. It may be especially effective against headache pain in veterans and individuals with severe trauma-related sleep issues (76).

In a prospective observational study, Ruff et al. (2009) evaluated whether treating impaired sleep using prazosin was associated with PPTH headache frequency and severity among OIF/OEF veterans with blast-exposure mTBI (n=74) (72). Of the 71 participants diagnosed with PTSD, 69 reported poor sleep. Prazosin use was associated with reduced headache intensity and frequency, more restful sleep, and improved cognitive performance. Addressing impaired sleep associated with PTSD may be an effective initial treatment strategy for headaches in OIF/OEF veterans with blast-induced mTBI and PTSD, but findings should be interpreted cautiously.

In an RCT, Mayer et al. (2023) evaluated the efficacy of prazosin for the prophylactic treatment of PPTH in active-duty service members and veterans (60) with blast or blunt mTBI (n=48). PTSD was present in 66% of the prazosin group and 81% of the placebo group. The prazosin group was associated with a significantly decreased incidence of headaches. Self-reported disability related to headaches was also significantly lower in the prazosin group relative to placebo. The study’s strengths included a rigorous RCT design, validated outcome measures, and an adequate study completion rate. However, the small sample size, long trial duration, minimal female representation, and relatively high prazosin doses (maximum dose of 5 mg morning and 20 mg evening) without prior dose-finding limit its broader applicability, and this RCT was assessed to have some concern of risk of bias.

#### Antidepressants and Serotonergic Pathways

Two studies investigated the efficacy of antidepressant agents as preventatives for PPTH. Antidepressants are thought to offer therapeutic benefits for headache symptoms (77), potentially reducing some of the comorbid affective burden. Mechanistically, it is thought that antidepressants may help reduce the frequency and severity of headaches by influencing serotonin and other neurotransmitters (78,79). Another study evaluated several types of medication whose theoretical efficacy may be associated with increased serotonin.

In an RCT, Hurwitz et al. (2020) evaluated the efficacy and safety of amitriptyline in preventing the development of PPTH after mTBI (n=50) on participants recruited from emergency departments, acute care admissions, outpatient clinics, and community brain injury events (61). Findings were not conclusive about the efficacy of amitriptyline, and the risk of bias was assessed as high due to several real-world challenges, including recruitment difficulties, medication adherence issues, and incomplete daily diary records. Amitriptyline may be effective for migraine prevention, but this phase II trial did not demonstrate efficacy against PPTH.

Using a retrospective cohort design, Cushman et al. (2019) evaluated the association of gabapentin and TCAs with headache symptoms over time after mTBI (66). Headache and symptom scores, which comprised 22 post-concussion related symptoms, improved over time for all patients (n=277), regardless of medication status, but piecewise regression analysis suggested that symptoms may have improved more rapidly with gabapentin or TCAs compared to the no-med group. However, the authors note that these changes could have been a result of natural recovery or a placebo effect remains unclear. Despite the large sample size and statistical rigor used to explore the treatment effect, the study was limited by its retrospective design, lack of a control group, with a substantial number of patients lacked follow-up data, raising concerns about potential differences in outcomes among excluded participants.

Erickson et al. (2011) conducted a retrospective observational study with follow-up to evaluate treatment outcomes in US Army soldiers (n=100) with PPTH attributable to mTBI (71). At baseline, 52% screened positive for PTSD and 38% for depression. Triptan class medications were often effective for aborting acute headache attacks, including both blast-related and non-blast-related PPTH. Topiramate is a medication thought to reduce electrical bursts associated with epilepsy, but it is also thought to help calm processes that stimulate migraines (80); it was associated with a significant reduction in headache frequency. However. Low-dose TCAs did not show a significant impact on headache frequency. A comprehensive headache management plan that addressed PTSD and sleep issues was associated with significantly lower headache-related disability. The study’s retrospective design, lack of a control group, multiple medication classes being evaluated, and focus on a military cohort limit generalizability and preclude definitive conclusions on treatment efficacy. However, we highlight the use of behavioral interventions (i.e., headache management and sleep plan) as potentially effective headache disability treatments.

#### Botulinum Toxins (Botox)

Three studies investigated the efficacy of botulinum toxins as a treatment for PTH/PPTH. Mechanistically, onabotulinumtoxinA (BoNT/A) is believed to exert its effects primarily through presynaptic inhibition of nociceptive neurotransmitters and neuropeptide release (e.g., glutamate, substance P, CGRP), leading to reduced neuroinflammation and muscle relaxation via blockade at the neuromuscular junction (64,81). Clinically, these mechanisms translate into meaningful reductions in headache burden, positioning BoNT/A as a valuable preventive treatment for PTH, especially in patients with limited response to conventional therapies (63).

In a retrospective, uncontrolled design, Yerry et al. (2015) provided preliminary data on the use of BoNT/A for PPTH in military service members (63). Participants (n=64) reported mTBI, including 56.3% with blast-related injuries. They reported that treatment with BoNT/A was associated with headache improvement, and slightly more than half of the cohort were able to maintain or return to active military service. These results should be interpreted as hypothesis-generating rather than confirmatory.

In an RCT, Zirovich et al. (2021) evaluated the efficacy of a single dose of BoNT/A in treating PPTH patients (n=40) reporting mixed complicated and uncomplicated mTBI (62). BoNT/A may be clinically effective in reducing headache frequency and pain severity in patients with PPTH. Methodological rigor and clinically meaningful outcomes were among the strengths of this study. However, there are some concerns of bias due to the small sample size, single-dose design, and short follow-up period.

Kazerooni et al. (2015) designed a retrospective case series to support the off-label use of incobotulinumtoxinA in reducing headache frequency and intensity among veterans with chronic migraine (n=21), including a subset with PPTH (n=6) following TBI (64). Among these individuals with PPTH, five reported clinically meaningful improvement, with a reduction in headache days per month from a mean of 19.5 to 6.7. Although incobotulinumtoxinA is not FDA-approved for chronic migraine, the findings suggest potential benefits for patients with PPTH, with the added benefits of lower cost and reduced drug wastage. However, the study’s retrospective design, small sample size, and lack of a control group limit the generalizability of the findings. Larger, prospective studies are needed to determine the efficacy of incobotulinumtoxinA in treating PPTH specifically.

#### Antidyskinetic

A single study investigated the efficacy of amantadine in the treatment of various post-concussion syndromes, including headaches. Amantadine hydrochloride is thought to enhance neural recovery after TBI through its dual mechanism as a dopamine receptor agonist and N-Methyl-D-aspartate (NMDA) receptor antagonist (82). Amantadine has shown promise in treating cognitive and behavioral symptoms following TBI (83), including reducing arousal symptoms and aggression and improving memory in TBI patients (84,85).

Using a retrospective observational design, Carabenciov et al. (2019) evaluated the effect of amantadine on post-concussion symptoms, particularly PPTH; 33 patients were reported by physician documentation to have PPTH (65). Amantadine may be a reasonable treatment option for post-concussion headaches, though further investigation is warranted.

### Abortive medications

#### Analgesics, NSAIDs, and Triptans

A single study investigated the efficacy of specific rescue agents as a treatment for PTH/PPTH. Current abortive treatments recommended for PTH work on mechanisms thought to influence primary headaches. To reduce headache pain, NSAIDs reduce inflammation and block pain signals (86), and triptans act on serotonin receptors to reduce dilation of blood vessels in the brain (87,88). Ditommaso et al. (2014) examined medication usage patterns for headache treatment 3, 6, or 12 months after mTBI using a prospective observational study (70). Over 70% of patients used over-the-counter analgesics such as acetaminophen and NSAIDs for headache control, with mixed results. Triptans were underutilized in this sample (8%); no conclusions can be drawn, but investigating triptans may be a path for future studies.

#### Anti-nausea (metoclopramide)

Two related studies investigated the efficacy of nausea medication as a treatment for acute PTH rescue. Metoclopramide, a dopamine receptor antagonist with analgesic properties, was not developed to treat headaches. However, it is often administered off-label as a first-line treatment in the ED setting due to its effectiveness in relieving both headache pain and associated nausea in acute migraine management (89). Metoclopramide has been shown to suppress central trigeminovascular nociception, suggesting an anti-neuroinflammation mechanism (90). Diphenhydramine is often co-administered in EDs to reduce the side effects of metoclopramide (91).

Friedman et al. (2018) conducted a prospective, open-label study of IV metoclopramide (plus diphenhydramine, M + D) for patients presenting to the ED with acute PTH (68). Seventy-one percent of patients (n=21) reported headache relief within 1 hour of treatment with M+D, and effectiveness increased to 95% by 2 hours. At 48 hours, 55% of patients maintained sustained headache relief without needing rescue medication. Effects were temporary: by one week, more than a quarter of patients still experienced frequent headaches, and 21% sought further medical care. Affective/mood changes were minimal. Despite the good tolerability of the medications, the design lacked rigor; there was no comparison group, and the sample was small with a short follow-up period.

Building on previous findings, Friedman et al. (2021) conducted a double-blind, placebo-controlled RCT to evaluate the efficacy of M + D for patients (n=160) experiencing acute PTH following head trauma presented in the ED (69). Similar to earlier results, M+D was associated with significant improvement in headache frequency and pain at one hour compared to placebo. There may have been a trend toward fewer mood-related symptoms in the treatment group compared to placebo, though these should be interpreted cautiously. In the short term, M+D may be recommended. Although the study was assessed as low risk of bias, its generalizability is limited by its urban ED setting; additionally, no long-term follow-up was reported.

#### Anti-inflammatory

A single study investigated the efficacy of antioxidant agents as a treatment for acute PTH following mTBI. N-acetyl-l-cysteine (NAC) has emerged as a promising therapeutic agent for mitigating post-concussive symptoms during the acute phase following TBI due to its potent antioxidant, anti-inflammatory, and neuroprotective properties (92,93).

Hoffer et al. (2013) implemented an RCT to evaluate the effect of NAC on the symptoms associated with blast exposure mTBI in a combat setting (n=81) (67) with 53 of them reporting having headaches. NAC, especially when initiated early (within 24 hours), was associated with faster symptom resolution by day seven post-blast; however, 24 participants continued to report unresolved headaches at that time. This was the first RCT conducted in an active combat zone and was assessed as having a low risk of bias, supported by its rigorous design and adequate sample size. However, limitations included a short follow-up period, a homogeneous military sample, and logistical challenges inherent to the combat environment.

## Discussion

This systematic review highlights the emerging landscape of pharmacological interventions for headaches following TBI. Beyond evaluating overall efficacy, we aimed to assess the extent to which these included studies addressed adverse effects, headache frequency, pain intensity, headache-related disability, and overall QoL. According to the effect direction summary, many studies reported positive effects. However, the few rigorously executed RCTs and controlled studies limit our ability to draw inferences.

Erenumab may demonstrate clinical benefits for both acute and persistent PTH by significantly reducing the frequency of moderate to severe headache days and improving QoL (57–59). MaB-derived therapies in general have become a cornerstone of the pharmaceutical industry (94). Since 2018, MaBs targeting CGRP activity have been employed in targeted migraine prophylaxis by reducing the neuroinflammation (95). An obvious next step was the evaluation of these therapies for PTH treatment. Individuals with PPTH demonstrate hypersensitivity to CGRP; CGRP infusions reliably induce migraine-like headaches in this population (74,96). Preclinical models supported that early and continuous blockade of CGRP using MaB-derived therapies after mTBI prevents both acute PTH and the establishment of a sensitized state that leads to PPTH (97). The potential role of CGRP in the pathogenesis of PTH makes CGRP antagonists like erenumab an attractive mechanism-based treatment approach.

Erenumab might be one of the most promising therapies available, but there is a conspicuous lack of RCTs or other controlled studies evaluating its effectiveness in this population. The included erenumab studies employed open-label and retrospective designs, and 2 of the 3 were notably underpowered. Additionally, injections may be a deterrent for some individuals—particularly those who prefer oral delivery or are hesitant about needles.

Among veterans and active-duty service members who report a high incidence of psychiatric comorbidities, prazosin is a potential therapeutic option that could reduce the frequency of headaches caused by TBI by improving sleep. Improved sleep may have additional behavioral benefits, improving QoL and reducing psychiatric distress (60). Only one RCT in the present review evaluated prazosin for PPTH, though off-label use in VA/DoD settings for PTSD-related sleep disturbances is common. The secondary benefits of improved sleep—particularly reduced PTSD symptoms—may contribute to its observed impact on headache reduction, suggesting a potential indirect pathway through behavioral regulation. Larger clinical trials are needed to confirm the extent of prazosin’s potential benefits among those with comorbid PPTH and psychiatric distress. Dose-finding studies would also help determine whether a lower dose could retain efficacy while decreasing the frequency of adverse events.

Treatment approaches for military populations must account for common comorbidities including PTSD—which affects nearly 30% of veterans and service members (98)—as well as high rates of anxiety, depression, and medication overuse (99). Among individuals with comorbid PPTH and affective distress, some pharmacologic interventions that target psychiatric and/or sleep-related conditions seem to be effective. A multidisciplinary care model that combines headache management medication with behavioral interventions may be especially effective (41).

Metoclopramide (plus diphenhydramine) relieved acute phase headache pain (68,69). Headache remains a common complaint among the 1.4 million patients who visit U.S. EDs annually after head trauma (100), and future research should explore optimal dosing and duration of this treatment and whether early intervention can improve longer-term outcomes, including comorbid symptoms like depression, sleep disturbances, and anxiety, which often affect individuals with PTH.

Overall, this review builds upon and extends prior work by offering a comprehensive and up-to-date synthesis of pharmacologic treatments for patients with acute PTH or PPTH. Consistent with prior reviews (34,101), our conclusions are limited by the availability of high-quality, powered trials evaluating both abortive and preventive pharmacologic treatments for both acute and persistent PTH. However, we believe that this work adds value to the body of research. First, it is necessary to periodically reassess the literature and reinterpret the data through the most recent lens. Second, while previous reviews incorporated many types of treatments and various populations (34), we focus exclusively on pharmacological interventions in adult populations to deliver a precise assessment of treatment effectiveness (98–101).

Use of pain medication as a first-line treatment option increases the risk of drug overuse and over-prescription, particularly when providers attempt to manage complex symptom profiles such as chronic pain, sleep disruption, and affective distress. Despite these obvious drawbacks, pharmacological treatments remain the most accessible pain management option, especially for individuals with limited resources or time. This review may assist healthcare providers by summarizing the most up-to-date, evidence-based therapeutic recommendations.

### Future Research Direction

There is a clear need for powered, randomized, placebo-controlled clinical trials to rigorously evaluate the efficacy and safety of both abortive and preventative treatments for acute phase PTH and PPTH. Current evidence is largely observational or based on expert opinion, with few RCTs. Further research is warranted to evaluate newer treatment options or re-evaluate old treatment paradigms across diverse populations and headache phenotypes.

It is important to recognize that, while the neuroinflammation model informs our understanding of the pathophysiology of primary headaches and has effectively provided key targets for pharmacotherapies, it may be an incomplete model to guide the development of treatments for secondary headaches. Injury mechanisms leading to PTH or PPTH—such as motor vehicle collisions, combat-related blast exposures, and physical assaults—are often accompanied by significant psychological stressors that can influence symptom expression, treatment response, and recovery trajectories. Investigators who explore pathophysiology and treatment models that account for both biological and psychological dimensions of PPTH may develop more consistently effective and personalized therapies.

### Strengths and Limitations

This systematic review provides a comprehensive synthesis of pharmacological treatments for acute phase PTH and PPTH in adults, incorporating both RCTs and observational studies across military and civilian populations. Key strengths include a registered protocol (PROSPERO), a librarian-assisted and PRESS-reviewed search strategy, adherence to PRISMA and SWiM guidelines, and rigorous risk-of-bias assessments using validated tools (RoB 2 and NOS). The review offers clinically relevant insights by focusing on adult populations and well-defined headache outcomes. However, the findings are limited by methodological heterogeneity among controlled studies, and the rest were retrospective or observational. Additionally, the exclusion of non-English and unpublished literature may have introduced bias. Despite these limitations, the review identifies important gaps and offers direction for future high-quality research. Finally, while we acknowledge that meta-analyses may often provide added insight, the substantial clinical and methodological heterogeneity across studies was assessed by the authors to be more likely to introduce additional bias than to answer questions, and so meta-analysis was not conducted.

## Conclusion

Pharmacological therapies are the first line of treatment for PTH/PPTH. Treatments developed for primary headache disorders have shown some effectiveness against PTH/PPTH pain, but there is currently no specific pharmacological therapy approved for it. Among individuals with PPTH and comorbid psychiatric distress, prazosin showed the best outcomes; among individuals with PPTH but no reported psychiatric distress, erenumab seemed to be most consistently effective. Among individuals with acute phase PTH, metoclopramide (plus diphenhydramine to suppress side effects) demonstrated short-term efficacy as a headache abortive in emergency care settings. This review also discussed the limits of the neuroinflammation model to explain functional differences in treatment outcomes between PTH/PPTH and primary headache disorders. Further research is needed to investigate the underlying differences between these headache types and to develop targeted pharmacological treatments for people with both acute and persistent PTH.

## Data Availability

All data used in this study are from previously published sources, which are cited in the manuscript.

## Abbreviations

TBI: Traumatic Brain Injury
mTBI: Mild Traumatic Brain Injury
PTH: Post-Traumatic Headache
PPTH: Persistent Post-Traumatic Headache
PTSD: Posttraumatic Stress Disorder
QoL: Quality of Life
RCT: Randomized Controlled Trial

## PubMed (554) (Appendix1)

(“Post-Traumatic Headache”[mesh] OR ((“Brain Injuries, Traumatic”[mesh] OR “whiplash injuries”[mesh] OR whiplash[tiab] OR “whip-lash”[tiab] OR “whip lash”[tiab] OR post traumatic[tiab] OR post-traumatic[tiab] OR posttraumatic[tiab] OR post concussion[tiab] OR post-concussion[tiab] OR postconcussion[tiab] OR “traumatic brain injury”[tiab] OR tbi[tiab] OR tbis[tiab] OR “Craniocerebral Trauma”[mesh] OR “Craniocerebral Trauma”[tiab]) AND (headache[mesh] OR “migraine disorders”[mesh] OR headache[tiab] OR “head ache” [tiab] OR migraine[tiab] OR cephalodynia*[tiab] OR cephalalgia*[tiab] OR cephalgia*[tiab] OR hemicrania[tiab])))

AND

(therapeutics[mesh] OR therapy[sh] OR treatment*[tiab] OR therap*[tiab] OR pharm*[tiab])

AND

(“randomized controlled trial” [pt] OR “controlled clinical trial“[pt] OR randomized[tiab] OR placebo[tiab] OR “drug therapy“[sh] OR randomly [tiab] OR trial[tiab] OR groups[tiab] OR “systematic review”[pt] OR “meta-analysis”[pt] OR “systematic review*“[tiab] OR “systematic literature review”[tiab] OR “integrative review“[tiab] OR “meta-analysis“[tiab] OR “meta analysis“[tiab])

NOT (“animals“[mesh] NOT (“animals“[mesh] AND “humans“[mesh]))

## CINAHL - 241

(((MH “Brain Injuries+”) OR (MH “whiplash injuries”) OR (TI whiplash OR AB whiplash) OR (TI “whip lash” OR AB “whip lash”) OR (TI “post traumatic” OR AB “post traumatic”) OR (TI posttraumatic OR AB posttraumatic) OR (TI “post concussion” OR AB “post concussion”) OR (TI postconcussion OR AB postconcussion) OR (TI “traumatic brain injury” OR AB “traumatic brain injury”) OR (TI tbi OR AB tbi) OR (TI tbis OR AB tbis) OR (TI “Craniocerebral Trauma” OR AB “Craniocerebral Trauma”) OR (MH “Head Injuries+”)) AND ((MH headache+) OR (MH “migraine+”) OR (TI headache OR AB headache) OR (TI “head ache” OR AB “head ache”) OR (TI migraine OR AB migraine) OR (TI cephalodynia* OR AB cephalodynia*) OR (TI cephalalgia* OR AB cephalalgia*) OR (TI cephalgia* OR AB cephalgia*) OR (TI hemicrania OR AB hemicrania)))

AND

((MH therapeutics+) OR (TI treatment* OR AB treatment*) OR (TI therap* OR AB therap*) OR (TI pharm* OR AB pharm*))

AND

((PT “randomized controlled trial”) OR (PT “controlled clinical trial”) OR (TI randomized OR AB randomized) OR (TI placebo OR AB placebo) OR “Drug Therapy” OR (TI randomly OR AB randomly) OR (TI trial OR AB trial) OR (TI groups OR AB groups) OR (PT “systematic review”) OR (PT meta-analysis) OR (TI “systematic review*” OR AB “systematic review*”) OR (TI “systematic literature review” OR AB “systematic literature review”) OR (TI “integrative review” OR AB “integrative review”) OR (TI meta-analysis OR AB meta-analysis) OR (TI “meta analysis” OR AB “meta analysis”))

NOT ((MH animals+) NOT ((MH animals+) AND (MH human+)))

## PsycINFO - 102

(((MH “ Traumatic Brain Injury+”) OR (TI “post traumatic” OR AB “post traumatic”) OR (TI posttraumatic OR AB posttraumatic) OR (TI “post concussion” OR AB “post concussion”) OR (TI postconcussion OR AB postconcussion) OR (TI “traumatic brain injury” OR AB “traumatic brain injury”) OR (TI tbi OR AB tbi) OR (TI tbis OR AB tbis) OR (TI “Craniocerebral Trauma” OR AB “Craniocerebral Trauma”) OR (MH “Head Injuries+”)) AND ((MH headache+) OR (MH “migraine headache+”) OR (TI headache OR AB headache) OR (TI “head ache” OR AB “head ache”) OR (TI migraine OR AB migraine) OR (TI cephalodynia* OR AB cephalodynia*) OR (TI cephalalgia* OR AB cephalalgia*) OR (TI cephalgia* OR AB cephalgia*) OR (TI hemicrania OR AB hemicrania))))

AND

((MH treatment+) OR (TI treatment* OR AB treatment*) OR (TI therap* OR AB therap*) OR (TI pharm* OR AB pharm*))

AND

NOT (MH animals)

## Scopus - 1159

(INDEXTERMS(“Post-Traumatic Headache”) OR ((INDEXTERMS(“Brain Injuries, Traumatic”) OR INDEXTERMS(“whiplash injuries”) OR TITLE-ABS(whiplash) OR TITLE-ABS(whip-lash) OR TITLE-ABS(“whip lash”) OR TITLE-ABS(“post traumatic”) OR TITLE-ABS(post-traumatic) OR TITLE-ABS(posttraumatic) OR TITLE-ABS(“post concussion”) OR TITLE-ABS(post-concussion) OR TITLE-ABS(postconcussion) OR TITLE-ABS(“traumatic brain injury”) OR TITLE-ABS(tbi) OR TITLE-ABS(tbis) OR INDEXTERMS(“Craniocerebral Trauma”) OR TITLE-ABS(“Craniocerebral Trauma”)) AND (INDEXTERMS(headache) OR INDEXTERMS(“migraine disorders”) OR TITLE-ABS(headache) OR TITLE-ABS(“head ache”) OR TITLE-ABS(migraine) OR TITLE-ABS(cephalodynia*) OR TITLE-ABS(cephalalgia*) OR TITLE-ABS(cephalgia*) OR TITLE-ABS(hemicrania))))

AND

(INDEXTERMS(therapeutics) OR “Therapy” OR TITLE-ABS(treatment*) OR TITLE-ABS(therap*) OR TITLE-ABS(pharm*))

AND

(DOCTYPE(“randomized controlled trial”) OR DOCTYPE(“controlled clinical trial”) OR TITLE-ABS(randomized) OR TITLE-ABS(placebo) OR “Drug Therapy” OR TITLE-ABS(randomly) OR TITLE-ABS(trial) OR TITLE-ABS(groups) OR DOCTYPE(“systematic review”) OR DOCTYPE(meta-analysis) OR TITLE-ABS(“systematic review*”) OR TITLE-ABS(“systematic literature review”) OR TITLE-ABS(“integrative review”) OR TITLE-ABS(meta-analysis) OR TITLE-ABS(“meta analysis”))

AND NOT (INDEXTERMS(animals) AND NOT (INDEXTERMS(animals) AND INDEXTERMS(humans)))

## Cochrane (136)

([mh “Post-Traumatic Headache”] OR (([mh “Brain Injuries, Traumatic”] OR [mh “whiplash injuries”] OR whiplash:ti,ab OR whip-lash:ti,ab OR “whip lash“:ti,ab OR “post traumatic“:ti,ab OR post-traumatic:ti,ab OR posttraumatic:ti,ab OR “post concussion“:ti,ab OR post-concussion:ti,ab OR postconcussion:ti,ab OR “traumatic brain injury“:ti,ab OR tbi:ti,ab OR tbis:ti,ab OR [mh “Craniocerebral Trauma”] OR “Craniocerebral Trauma“:ti,ab) AND ([mh headache] OR [mh “migraine disorders”] OR headache:ti,ab OR “head ache“:ti,ab OR migraine:ti,ab OR cephalodynia*:ti,ab OR cephalalgia*:ti,ab OR cephalgia*:ti,ab OR hemicrania:ti,ab)))

AND

([mh therapeutics] OR [mh /TH] OR treatment*:ti,ab OR therap*:ti,ab OR pharm*:ti,ab) AND

(“randomized controlled trial“:pt OR “controlled clinical trial“:pt OR randomized:ti,ab OR placebo:ti,ab OR [mh /DT] OR randomly:ti,ab OR trial:ti,ab OR groups:ti,ab OR “systematic review“:pt OR meta-analysis:pt OR (“systematic” NEXT review*):ti,ab OR “systematic literature review“:ti,ab OR “integrative review“:ti,ab OR meta-analysis:ti,ab OR “meta analysis“:ti,ab)

NOT ([mh animals] NOT ([mh animals] AND [mh humans]))

